# Neonatal Stroke Active Surveillance Study in the United Kingdom and Ireland with meta-analysis of surveillance studies

**DOI:** 10.1101/2025.09.22.25336405

**Authors:** T’ng Chang Kwok, Robert A Dineen, William P Whitehouse, Richard M Lynn, Niamh McSweeney, Don Sharkey, the UK/Irish Neonatal Stroke Study Collaborative

## Abstract

**Background:** Neonatal stroke is a rare but devastating condition. This study explored the incidence, clinical presentation, management and short-term outcomes of neonatal stroke across five countries in the United Kingdom (UK) and Republic of Ireland (ROI) in the post-COVID-19 era.

**Methods:** This active surveillance study identified neonatal stroke cases presenting before 90 days of life between March 2022-April 2023 in the UK and ROI using the monthly British Paediatric Surveillance Unit (BPSU) reporting system. Reporting clinicians completed questionnaires and uploaded de-identified neuroimages via a purpose-built data platform. A meta-analysis of neonatal stroke active surveillance studies was performed.

**Results:** 68 neonatal stroke cases were reported. UK incidence of neonatal stroke was 9·0 per 100,000 live births (95% CI 6·9-11·6). Three-quarters of the cases were arterial ischaemic and unilateral. Arterial ischaemic and cerebral venous sinus thrombosis (CVST) strokes commonly presented with seizures at two-three days of age, while haemorrhagic stroke commonly presented with encephalopathy in the first ten days of age. Neonatal stroke cases were associated with fetal distress *in utero*. 61% and 28% of infants had an umbilical cord pH <7·25 and required significant resuscitation at birth, respectively. A meta-analysis of 3,607,864 infants found the incidence and associated conditions were similar to pre-COVID-19 surveillance studies in Germany and Australia.

Neonatal stroke guidelines were available in a quarter of the reporting hospitals. 87% of infants with arterial ischaemic and CVST stroke received antiseizure medications. 82% of infants were discharged home at a median of 12 days old with antiseizure medications (42%) alongside paediatric/neonatal (91%), physiotherapy (77%) and paediatric neurology (63%) follow-up.

**Conclusions:** Neonatal stroke is a rare disease with distinct subtypes associated with different clinical presentations, timings and management strategies, highlighting the need to better understand this condition. The incidence pre- and post-COVID-19 appeared similar.

**Funding:** 2019 BPSU Sir Peter Tizard Award

## Introduction

Neonatal stroke is a rare but devastating condition that often results in lifelong brain injury and impairment in children across the globe.^1^ It is associated with significant healthcare costs^2^ and impact on education services and parents^3^ to support surviving infants. Although stroke is more common in newborn infants than in older children,^4^ the true contemporary incidence of neonatal stroke remains unclear, especially after the COVID-19 pandemic. There is significant heterogeneity in the reported incidence rate of neonatal stroke, especially in Europe, ranging from 6.1 to 47.3 per 100,000 live births,^4^ making the estimation of the true burden of neonatal stroke challenging. Robust, prospective population-based studies reporting clinical management and outcomes of neonatal stroke are lacking.

Neonatal stroke differs from stroke in older children and adults^5^ due to the immaturity and plasticity of the brain in newborn infants.^6^ As neonatal stroke is a rare condition, most clinicians may only manage a few cases throughout their career. This results in a poorly described condition with little recent knowledge on management and outcomes, unlike adult stroke. The lack of awareness of neonatal stroke may lead to delays in diagnosis and variation in practice in managing infants with neonatal stroke^7^. Using the established British Paediatric Surveillance Unit (BPSU; https://www.bpsu.org.uk/) for secure data collection, we undertook an active surveillance study to estimate the incidence of neonatal stroke across five countries in the United Kingdom (UK) and the Republic of Ireland (ROI). Our secondary aims were to analyse the proportion of the different types of neonatal stroke, alongside the predisposing factors, clinical presentation, management, and short-term outcomes of neonatal stroke. Thirdly, a literature search with meta-analysis of neonatal stroke incidence and associated conditions was performed to compare our study results with findings from previous active surveillance studies. This provided the opportunity to compare the neonatal stroke incidence pre-and post-COVID-19 pandemic.

## Methods

### Study design

The full protocol for this study has been published previously^8^. This is the first multinational prospective cohort study using a new purpose-built integrated case notification-data collection platform and safe haven based on the standard BPSU active surveillance approach^9^. The study received ethics approval from the UK Health Research Authority Research Ethics Committee (Reference 21/EM/0110) on advice from the Confidentiality Advisory Group (England and Wales) (Reference 21/CAG/0061); the Public Benefit and Privacy Panel (Scotland) (Reference 2122-0006 Kwok); and the chair of the Privacy Advisory Committee (Northern Ireland). Results are reported in accordance with Strengthening the Reporting of Observational Studies in Epidemiology (STROBE) guidelines^10^.

### Procedures / Active surveillance

From 1 April 2022, paediatricians across the UK and ROI received a monthly BPSU electronic reporting card to report active surveillance studies. If they had seen a case of neonatal stroke, they would be directed to the surveillance data collection platform to notify and complete an electronic data collection form for each case seen between 1^st^ March 2022 and 31^st^ April 2023 that fulfilled the case definition. The data collection form contained information on the pregnancy, clinical presentation, investigation and treatment received, as well as the short-term outcome following the initial hospital admission. De-identified neuroimages were uploaded to the data collection platform, where possible. Paediatricians also confirmed on the monthly reporting card if no neonatal stroke cases were seen in the preceding month (active negative surveillance).^9^ To increase awareness and reporting, the study was also publicised via the British Association of Perinatal Medicine and the British Paediatric Neurology Association, the two leading national bodies whose members were most likely to see cases.

### Participants / Case definition

The neonatal stroke case was defined as any infant presenting in the first 90 days of age with any neurological symptoms and either neuroimaging or neuropathology demonstrating disruption of cerebral blood flow. Infants with hypoxic ischaemic encephalopathy, germinal matrix/intraventricular haemorrhage in preterm infants, metabolic injury, encephalitis, accidental/non-accidental injury, vascular anomalies, and brain tumours were excluded from the case definition. The case definition was based on international consensus and previously published studies.^11,12^ The age range was extended to 90 days to capture infants that presented late after the first 28 days of age with neuroimaging or neuropathology suggesting stroke occurring in the neonatal period. The neuroimaging reports and/or neuroimages themselves were assessed by the research team (TK/NM and RD, respectively) to ensure that the reported case fulfilled the case definition. A consensus decision from the whole research team (TK, RD, WW and DS) was used for any cases which were unclear whether they met the case definition.

### Statistical analysis

Descriptive statistics were presented as numbers with percentages and median with interquartile ranges for categorical and continuous data, respectively. Cells with fewer than five infants were suppressed to avoid data disclosure that could be used to identify individual infants. The estimation of the incidence rate of neonatal stroke was only possible for the UK, as there was incomplete coverage of cases reported from the ROI. The first and last months of surveillance were excluded from the estimation of the incidence rate, in line with the standard BPSU approach.^9^ Live birth data from the Office for National Statistics,^13^ National Records of Scotland,^14^ and Northern Ireland Statistics and Research Agency^15^ were used as the denominators for the incidence estimation. The 95% confidence interval (CI) of the incidence was determined using the binomial exact method. Sensitivity analyses of the incidence rate were performed by excluding cases presented after the first 28 days of age and preterm infants. The incidence of neonatal stroke based on sex and ethnicity was derived from live birth data in 2022 rather than exact months. Ethnicity was based on the 2011 Office for National Statistics for UK-wide data collection^16^. Northern Ireland was excluded from the incidence rate determination by ethnicity due to a lack of live birth data by ethnicity.

### Meta-analysis

A literature search was performed on 29/7/25 in PubMed to identify active surveillance studies of neonatal stroke. Predisposing factors, clinical presentation and management of neonatal stroke were extracted if available. Meta-analysis of the incidence rate and associated conditions of neonatal stroke and its subtype was performed using the random-effects method and the *metaprop* package^17^ in STATA SE18.

### Patient and public involvement

The study was designed in partnership with Bliss, the national charity for families of infants born sick or premature and lay representatives of the BPSU scientific committee.

## Results

An average of 68% of 4,231 paediatricians returned the monthly BPSU reporting card (**Table S1**). 119 potentially eligible neonatal stroke cases were notified through the BPSU surveillance system. 25 (21%) notified cases were either lost to follow-up, or no response was received from notifying paediatricians. From the 94 data collection forms received, 21 (22%) provided neuroimages (**Figure 1**), while 55 (59%) provided the neuroimaging report. After excluding ineligible and duplicate cases, 68 cases met the neonatal stroke case definition (**Figure 2**). Of the 68 neonatal stroke cases, 53 (78%), 9 (13%), and 6 (9%) cases were arterial ischaemic, cerebral venous sinus thrombosis (CVST) and haemorrhagic stroke, respectively. Less than five and seven (10%) of the neonatal cases presented after the first 28 days of age or in preterm infants, respectively. 48 out of 67 cases (72%) were diagnosed during a neonatal unit admission.

**Figure 1.**
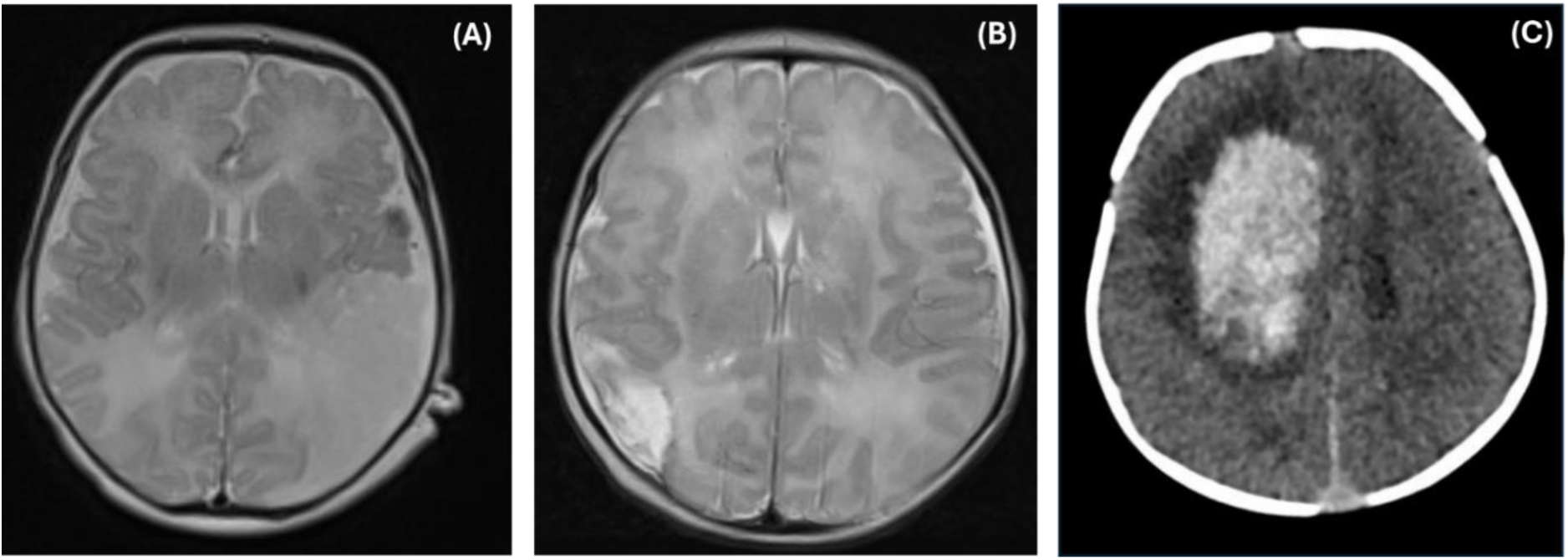
Example neuroimages submitted for review by the research team from three different infants. (A) Axial T2-weighted magnetic resonance imaging (MRI) scan showing acute left middle cerebral artery infarction; (B) Axial T2-weighted MRI scan showing subacute cortical infarction in the right middle cerebral artery territory; and (C) Plain computerised tomography (CT) scan demonstrating right frontal haemorrhagic stroke.

**Figure 2.**
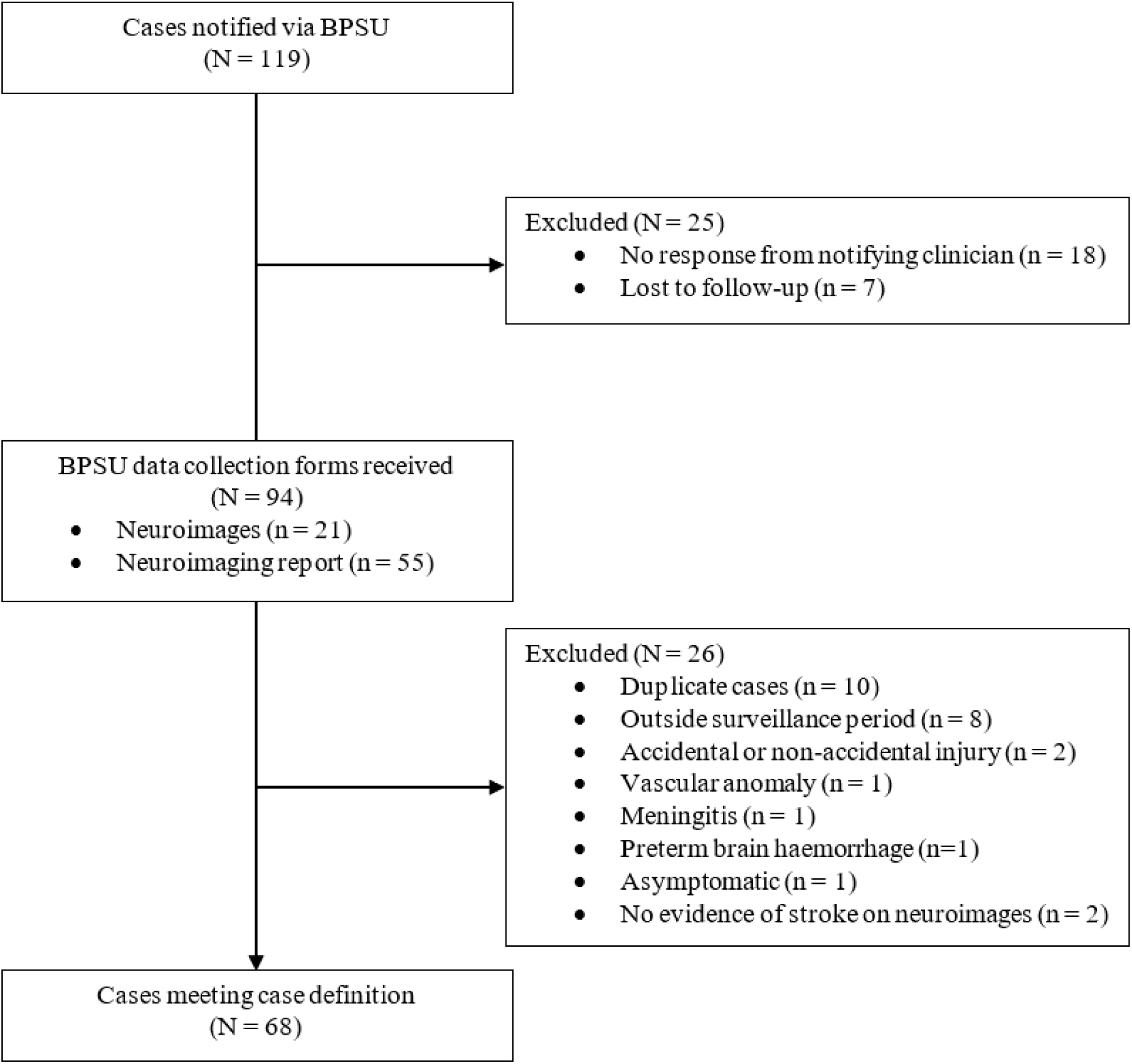
Flowchart of the case selection during the study period 1^st^ March 2022 to 31^st^ April 2023.

### Incidence

There were 668,247 live births and 60 neonatal stroke cases between 1 April 2022 to 31 March 2023 in the UK. Therefore, the overall incidence of neonatal stroke was 9·0 cases per 100,000 live births (95% CI 6·9 to 11·6 per 100,000 live births). The incidence of the individual type of neonatal stroke over the same study period was 6·9 (95% CI 5·0 to 9·2), 1·4 (95% CI 0·6 to 2·6), and 0·7 (95% CI 0·2 to 1·8) cases per 100,000 live births for arterial ischaemic, CVST and haemorrhagic stroke respectively. After excluding preterm infants and cases presenting after 28 days of age, the overall incidence was 8·2 cases per 100,000 live births (95% CI of 6·2 to 10·7). The incidence of neonatal stroke was higher in male (9·8 (95% CI 6·8 – 13·8)), white (10·1 (95% CI 7·3 – 13·5)) and Asian (12·2 (95% CI 5·8 – 22·4)) infants (**Table 1**).

**Table 1.** Incidence and demographics of neonatal stroke and its subtype in the United Kingdom from April 2022 to March 2023. *Live birth data for the year 2022 was used as the denominator rather than the individual months. ^†^Northern Ireland data was excluded due to a lack of live birth data by ethnicity. ^‡^Asian refers to Indian, Pakistani, Bangladeshi and Chinese. ^§^Missing data (n, %) for the following variables: gestational age (1, 2%) and birth weight (1, 2%).

| Incidence rate (95% confidence interval), per 100,000 live births |  |  |  |  |
| --- | --- | --- | --- | --- |
| Cohort | Neonatal stroke | Arterial ischaemic | Cerebral venous sinus thrombosis | Haemorrhagic |
| **Overall cohort** | 9.0 (6.9 - 11.6) | 6.9 (5.0 - 9.2) | 1.4 (0.6 - 2.6) | 0.7 (0.2 - 1.8) |
| Sex<sup>*</sup> |  |  |  |  |
| Male | 9.8 (6.8 - 13.8) | 7.5 (4.9 - 11.0) | 1.2 (0.3 - 3.0) | 1.2 (0.3 - 3.0) |
| Female | 7.9 (5.2 - 11.6) | 6.1 (3.7 - 9.4) | 1.5 (0.5 - 3.6) | 0.3 (0.0 - 1.7) |
| Ethnicity<sup>*,†</sup> |  |  |  |  |
| White | 10.1 (7.3 - 13.5) | 8.0 (5.6 - 11.1) | 1.1 (0.4 - 2.6) | 0.9 (0.2 - 2.3) |
| Asian / Asian British<sup>‡</sup> | 12.2 (5.8 - 22.4) | 8.5 (3.4 - 17.5) | 2.4 (0.3 - 8.8) | 1.2 (0.0 - 6.7) |
| Black / African / Caribbean / Black British | 5.8 (0.7 - 21.0) | 2.9 (0.1 - 16.2) | 2.9 (0.1 - 16.2) | N/A |
| Mixed / Multiple Ethnic Groups | 4.7 (0.6 - 16.9) | 2.3 (0.1 - 13.0) | 2.3 (0.1 - 13.0) | N/A |
| **Exclude preterm & present more than 28 days old** | 8.2 (6.2 - 10.7) | 6.4 (4.7 - 8.7) | 1.3 (0.6 - 2.6) | 0.4 (0.1 - 1.3) |
| Demographics, median (IQR) |  |  |  |  |
| Demographics | Overall neonatal stroke<br>(N = 68) | Arterial ischaemic<br>(N = 53) | Cerebral venous sinus thrombosis<br>(N = 9) | Haemorrhagic<br>(N = 6) |
| Gestational age at birth (weeks)<sup>§</sup> | 39<sup>+5</sup> (38<sup>+2</sup> - 40<sup>+6</sup>) | 39<sup>+5</sup> (38<sup>+2</sup> - 40<sup>+6</sup>) | 40<sup>+3</sup> (39<sup>+1</sup> - 41<sup>+0</sup>) | 39<sup>+4</sup> (38<sup>+0</sup> - 41<sup>+0</sup>) |
| Birth weight (g)<sup>§</sup> | 3240<br>(2980 - 3650) | 3289<br>(2940 - 3580) | 3205<br>(3117.5 - 3442.5) | 3515<br>(3225 - 4100) |
| Age at presentation (days) | 2 (1 - 4) | 2 (1 - 3) | 3 (1 - 5) | 10 (3 - 34) |

### Case characteristics

#### Infant demographics

Most of the neonatal stroke cases occurred in term (90%), male (56%), and White (79%) infants (**Table 1**). The infant demographics were fairly similar across the different neonatal stroke types.

#### Antenatal / Perinatal factors

Mothers of infants with neonatal stroke were mainly primigravida (55%). 21% and 19% mothers with neonatal stroke infants had meconium-stained liquor and maternal diabetes, respectively. 52% infants with neonatal stroke were born vaginally, with 61% and 28% infants having a cord pH below 7·25 and requiring significant resuscitation at birth (defined as more than 1 set of inflation breaths), respectively. The remaining factors are described in **Table 2**.

**Table 2:** Table describing the demographics of the 68 neonatal stroke cases split by type of neonatal stroke. Cells with fewer than five infants were suppressed. Missing data (n, %) for the following variables: ethnicity (1, 2%), diagnosed in neonatal unit (1, 2%), focal neurology (4, 6%), abnormal tone (4, 6%), lethargy (5, 7%), poor feeding (6, 9%), apnoea (2, 3%), mode of delivery (4, 6%), Vitamin K (7, 10%), Apgar at 5 minutes (10, 15%), Apgar at 10 minutes (11, 16%), cord pH (32, 47%), significant resuscitation (1, 1.5%), multiple pregnancy (2, 3%), primigravida (6, 9%). pre-eclampsia (7, 10%), meconium (5, 7%), maternal thrombophilia/prothrombotic (10, 15%), maternal smoking (9, 13%), maternal diabetes (6, 9%), maternal fever (6, 9%), prolonged rupture of membrane (7, 10%), maternal substance abuse (6, 9%), family history of stroke (10, 15%), sepsis (2, 3%), congenital cardiac defect (2, 3%), and coagulation defect (17, 25%).

| Clinical characteristics | Overall cases<br>(N = 68) |
| --- | --- |
| **<u>Infant demographics</u>** |  |
| Gestational age at birth (weeks), median (IQR) | 39<sup>+5</sup> (38<sup>+2</sup> - 40<sup>+6</sup>) |
| Birth weight (g), median (IQR) | 3240 (2980 - 3650) |
| Male sex, n/N (%) | 38/68 (56) |
| Ethnicity, n/N (%) |  |
| White | 53/67 (79) |
| Asian / Asian British | 10/67 (14) |
| Black / African / Caribbean / Black British | <5 |
| Mixed / Multiple Ethnic Groups | <5 |
| **<u>Perinatal factors</u>** |  |
| Delivery mode, n/N (%) |  |
| Vaginal (spontaneous) | 19/68 (30) |
| Vaginal (instrumental) | 14/68 (22) |
| Emergency Caesarean | 22/68 (34) |
| Elective Caesarean | 9/68 (14) |
| Vitamin K at birth, n/N (%) | 61/61 (100) |
| Apgar scores, median (IQR) |  |
| 5 minutes | 9 (6 - 9) |
| 10 minutes | 9 (8 - 10) |
| Cord pH < 7.25, n/N (%) | 22/36 (61) |
| Significant resuscitation at birth, n/N (%) | 19/67 (28) |
| **<u>Maternal factors</u>** |  |
| Multiple pregnancy, n/N (%) | <5 |
| Primigravida, n/N (%) | 34/62 (55) |
| Pre-eclampsia, n/N (%) | 6/61 (10) |
| Meconium-stained liquor, n/N (%) | 13/63 (21) |
| Maternal thrombophilia/prothrombotic, n/N (%) | <5 |
| Maternal smoking, n/N (%) | <5 |
| Maternal diabetes, n/N (%) | 12/62 (19) |
| Maternal fever, n/N (%) | 8/62 (13) |
| Prolonged rupture of membrane, n/N (%) | 8/61 (13) |
| Maternal substance misuse, n/N (%) | <5 |
| Family history of neonatal stroke, n/N (%) | 0/58 (0) |
| **<u>Clinical presentation</u>** |  |
| Site of stroke, n/N (%) |  |
| Left | 33/68 (49) |
| Right | 19/68 (28) |
| Bilateral | 16/68 (24) |
| Age at presentation (days), median (IQR) | 2 (1 - 4) |
| Diagnosed in a neonatal unit, n/N (%) | 48/67 (72) |
| Signs, n/N (%) |  |
| Seizure | 61/68 (90) |
| Focal neurology | 18/64 (28) |
| Abnormal tone | 22/64 (34) |
| Decreased tone | 16/22 (73) |
| Lethargy / Altered consciousness | 25/63 (40) |
| Poor feeding | 25/62 (40) |
| Apnoea/bradycardia/desaturations | 32/66 (48) |
| Co-morbidities, n/N (%) |  |
| Sepsis | 8/66 (12) |
| Congenital cardiac defect | 12/66 (18) |
| Atrial septal defect / patent foramen ovale | 5/12 (42) |
| Ventricular septal defect | <5 |
| Transposition of great arteries | <5 |
| Pulmonary branch stenosis | <5 |
| Coagulation defect | <5 |
| Others |  |
| Congenital airway/pulmonary abnormalities | <5 |
| Hypoglycaemia | <5 |
| Hypopituitarism | <5 |
| Extracorporeal membrane oxygenation | <5 |

### Clinical presentation

Infants with neonatal stroke presented at a median age of 2 days. Arterial ischaemic and CVST strokes commonly presented at a median of two to three days of age, while haemorrhagic stroke commonly presented at a median of ten days of age. 52 (76%) infants had unilateral stroke. The three most common presenting signs were seizure, apnoea/bradycardia/desaturations and poor feeding, occurring in 90%, 48% and 40% of infants with neonatal stroke, respectively. Infants with arterial ischaemic and CVST stroke in our cohort had similar clinical presentations. Infants with haemorrhagic stroke commonly presented with poor feeding and altered consciousness. 18% infants with neonatal stroke had congenital cardiac defects (**Table 2**).

### Clinical management

Neonatal stroke guidelines were only available in 13 (25%) out of 51 hospitals which reported at least one case of neonatal stroke. All infants had neuroimaging to confirm the neonatal stroke diagnosis; 96% infants underwent MRI, and 4% infants underwent computed tomography (CT) scan. Electrocardiogram, cerebral function monitoring, cerebrospinal fluid culture, and echocardiogram were commonly performed in infants with neonatal stroke alongside blood tests of full blood count and coagulation screen. Genetic study and placental histology were performed in 23% and 18% neonatal stroke cases (**Table 3**).

**Table 3:** Table describing the clinical management and outcome of the 68 neonatal stroke cases. MRI = Magnetic Resonance Imaging. CT = Computed Tomography Missing data (n (%%) for the following variables: electroencephalogram (3, 4%), cerebral function monitoring (5, 7%), full blood count (1, 2%), coagulation screen (1, 2%), cerebral spinal fluid culture (1, 2%), genetic study (2, 3%), placental histology (23, 34%), therapeutic hypothermia (2, 3%), age at discharge (1, 2%), antiseizure medication on discharge (1, 2%), anticoagulant on discharge (5, 9%), aspirin on discharge (5, 9%), paediatric neurology follow-up (5, 9%), neonatal/general paediatric follow-up (2, 4%), community paediatrics follow-up (8, 14%), and physiotherapy follow-up (5, 9%).

| Clinical management/outcome | Overall cases<br>(N = 68) |
| --- | --- |
| ***Investigation*** |  |
| Neuroimaging, n/N (%) |  |
| MRI (no contrast) | 46/68 (68) |
| MRI (contrast) | 19/68 (28) |
| CT (no contrast) | 3/68 (4) |
| Electroencephalogram, n/N (%) | 43/65 (66) |
| Cerebral function monitoring, n/N (%) | 47/63 (75) |
| Echocardiogram, n/N (%) | 45/68 (66) |
| Full blood count, n/N (%) | 66/67 (99) |
| Coagulation screen, n/N (%) | 52/67 (78) |
| Cerebrospinal fluid culture, n/N (%) | 40/67 (60) |
| Genetic study, n/N (%) | 15/66 (23) |
| Placental histology, n/N (%) | 8/45 (18) |
| ***Treatment*** |  |
| Antiseizure medications, n/N (%) | 55/68 (81) |
| Phenobarbitone | 49/55 (89) |
| Levetiracetam | 27/55 (49) |
| Midazolam | 9/55 (16) |
| Phenytoin | 7/55 (13) |
| Lidocaine | <5 |
| Multiple antiseizure medications | 25/55 (45) |
| Anticoagulant, n/N (%) | 9/68 (13) |
| Heparin | 6/9 (67) |
| Enoxaparin/Tinzaparin/Rivaroxaban | <5 |
| Aspirin, n/N (%) | <5 |
| Tranexamic acid, n/N (%) | <5 |
| Therapeutic hypothermia, n/N (%) | 5/66 (8) |
| ***Discharge*** |  |
| Discharge home, n/N (%) | 56/68 (82) |
| Age at discharge (day), median (IQR) | 12 (9 - 16) |
| ***Medication on discharge*** |  |
| Antiseizure medication | 23/55 (42) |
| Anticoagulant | <5 |
| Aspirin | <5 |
| ***Follow-up*** |  |
| Paediatric neurologist / special interest<br>neurology | 32/51 (63) |
| Neonatal/General Paediatric | 49/54 (91) |
| Community Paediatric | 18/48 (38) |
| Physiotherapist | 39//51 (76) |
| Paediatric endocrinologist | <5 |
| Paediatric ophthalmologist | <5 |
| Paediatric cardiologist | <5 |
| Haematologist | <5 |

The majority of infants with neonatal stroke received antiseizure medications (81%), with phenobarbitone (89%) being the most commonly used, and nearly half (45%) received multiple types of antiseizure medications. Anticoagulant and aspirin were given to 13% and <5 cases, respectively, with therapeutic hypothermia provided in 8% of cases (**Table 3**).

### Outcome

56 (82%) infants were discharged home at a median age of 12 days old, with the remaining 12 infants still in hospital when the questionnaire was completed. Nearly half of the infants discharged home were discharged on antiseizure medications (42%), with <5 infants discharged on anticoagulants and aspirin. The majority of infants were discharged with neonatal/general paediatric (91%), physiotherapy (76%) and paediatric neurology (63%) follow-up. 38% infants who were discharged home were followed up by community paediatricians (**Table 3**).

### Meta-analysis

The literature search identified two further surveillance studies carried out in Germany (2,314,617 live births during the surveillance period)^18–20^ and Australia (around 625,000 live births during the surveillance period)^21,22^, respectively. Both studies were carried out in the five-year period prior to the COVID-19 pandemic. The reported incidences of neonatal stroke and its subtypes across the two surveillance studies were similar to those found in our study, except for haemorrhagic stroke, where our study reported a lower incidence (**Figure 3**). For the meta-analysis of the incidence rate, only term infants presenting with neonatal stroke in the first 28 days in our study were included to ensure consistency in the case definition across the surveillance studies. The associated factors and clinical presentations in our study were similar to the two surveillance studies (**Table 4, Table S2**).

**Figure 3.**
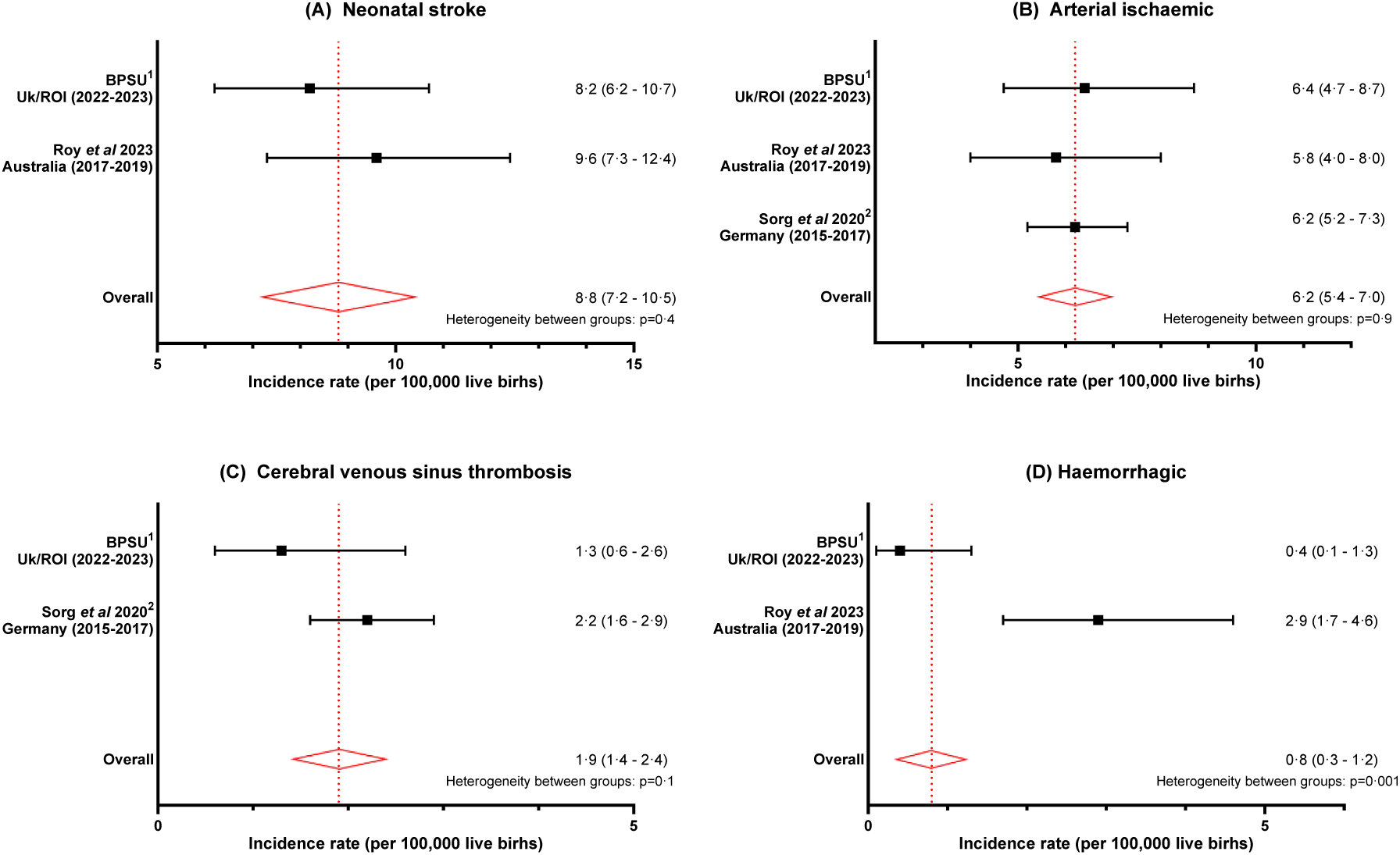
Forest plot of the incidence of neonatal stroke and its subtypes across the three active surveillance studies in the United Kingdom/Republic of Ireland (current study), Germany^18–20^ and Australia^21,22^. ^1^ Only term infants presenting in the first 28 days of life were included to ensure consistency in the case definition across the surveillance studies. ^2^ Live birth rates for 2015 to 2017 in Germany were determined from Statistisches Bundesamt Deutschland^23^.

**Table 4:** Meta-analysis of the percentage of cases (95% confidence interval) with the associated conditions for each of the three neonatal stroke subtypes across the three active surveillance studies in the United Kingdom/Republic of Ireland (current study), Germany^18–20^ and Australia^21,22^. N/A = Meta-analysis was not possible as data were not available from at least two studies.

| Characteristics,<br>% (95% confidence interval) | Arterial ischaemia |  | Venous thrombosis |  | Haemorrhage |  |
| --- | --- | --- | --- | --- | --- | --- |
|  | Pooled | Number of infants | Pooled | Number of infants | Pooled | Number of infants |
| **<u>Infant demographics</u>** |  |  |  |  |  |  |
| Preterm | 12.2 (7.6 - 16.8) | 193 | 21.9 (11.4 - 34.3) | 57 | N/A | N/A |
| Small for gestational age (<10th centile) | 17.5 (12.5 - 22.4) | 224 | 12.7 (3.9 - 21.5) | 55 | 7.3 (0 - 25.0) | 21 |
| Male sex | 63.7 (57.5 - 69.9) | 233 | 63.6 (51.6 - 75.6) | 60 | 58.6 (38.9 - 78.2) | 24 |
| White ethnicity | 85.9 (72.2 - 99.6) | 227 | 95.3 (90.0 - 100) | 59 | 73.2 (54.5 - 91.8) | 21 |
| **<u>Maternal risk factor</u>** |  |  |  |  |  |  |
| Maternal smoking | 3.1 (0 - 6.8) | 79 | N/A | N/A | 7.2 (0 - 17.7) | 23 |
| Maternal substance misuse | 4.3 (0 - 8.6) | 83 | N/A | N/A | 0 (0 - 7.3) | 23 |
| Family history of neonatal stroke | 0 (0 - 9.7) | 81 | N/A | N/A | 0 (0 - 7.2) | 22 |
| **<u>Pregnancy risk factors</u>** |  |  |  |  |  |  |
| Multiple pregnancy | 4.5 (1.8 - 7.1) | 230 | 6.5 (0.2 - 12.7) | 59 | 0.8 (0 - 12.1) | 24 |
| Primigravida | 56.5 (50.0 - 62.9) | 225 | 51.1 (38.5 - 63.8) | 58 | 56.6 (36.4 - 76.8) | 23 |
| Pre-eclampsia | 6.3 (3.1 - 9.5) | 218 | 11.5 (3.2 - 19.8) | 56 | 2.9 (0 - 16.8) | 23 |
| Prolonged rupture of membrane | 5.5 (0.6 - 10.4) | 81 | N/A | 59 | 0 (0 - 6.8) | 24 |
| Gestational diabetes | 10.4 (5.8 - 15.0) | 169 | N/A | N/A | N/A | N/A |
| Chorioamnionitis | 5.7 (2.3 - 9.2) | 174 | N/A | N/A | N/A | N/A |
| **<u>Perinatal risk factors</u>** |  |  |  |  |  |  |
| Mode of delivery |  |  |  |  |  |  |
| Spontaneous vaginal delivery | 26.9 (14.5 - 39.2) | 225 | 35.5 (23.2 - 47.9) | 53 | 47.8 (27.4 - 68.2) | 23 |
| Instrumental-assisted vaginal delivery | 15.5 (9.9 - 21.1) | 225 | 11.3 (2.8 - 19.8) | 53 | 25.7 (7.9 - 43.5) | 23 |
| Elective Caesarean section | 17.2 (12.3 - 22.2) | 225 | 7.6 (1.1 - 17.4) | 53 | 1.0 (0 - 12.9) | 23 |
| Emergency Caesarean section | 37.5 (28.5 - 46.6) | 225 | 41.9 (28.9 - 54.9) | 53 | 18.3 (3.7 - 38.3) | 23 |
| Cord pH <7.25 | 48.1 (35.5 - 60.7) | 58 | N/A | 48 | 36.9 (11.0 - 66.3) | 16 |
| Significant resuscitation | 31.4 (16.4 - 46.3) | 231 | 40.7 (28.1 - 53.2) | 59 | 35.3 (16.7 - 53.8) | 23 |
| Apgar <7 at 5 minutes | 17.0 (7.2 - 26.7) | 220 | 28.0 (15.6 - 40.4) | 50 | 15.7 (1.8 - 36.2) | 21 |
| Apgar <7 at 10 minutes | 7.0 (0.4 - 13.6) | 57 | N/A | N/A | 17.6 (0 - 35.7) | 17 |
| Meconium-stained liquor | 28.1 (18.6 - 37.7) | 84 | N/A | N/A | 2.7 (0 - 16.1) | 24 |
| **<u>Infant co-morbidity</u>** |  |  |  |  |  |  |
| Sepsis | 14.2 (5.1 - 23.3) | 218 | 15.2 (6.0 - 24.3) | 59 | 8.1 (0 - 27.3) | 19 |
| Congenital cardiac defect | 22.8 (13.3 - 32.2) | 71 | N/A | N/A | 33.3 (6.7 - 60.0) | 12 |
| Hypoglycaemia | 5.4 (2.4 - 8.4) | 217 | 8.2 (1.6 - 17.7) | 59 | 3.7 (0 - 20.3) | 19 |
| Received therapeutic hypothermia | 4.8 (1.8 - 7.8) | 195 | 13.3 (4.7 - 21.8) | 60 | N/A | N/A |
| **<u>Clinical presentation</u>** |  |  |  |  |  |  |
| Seizure | 75.7 (67.9 - 82.4) | 197 | 76.0 (63.6 - 86.6) | 60 | N/A | N/A |
| Hypotonia | 17.4 (12.1 - 22.8) | 194 | 31.8 (20.0 - 43.5) | 59 | N/A | N/A |
| Lethargy | 6.0 (2.9 - 9.2) | 193 | 23.7 (12.9 - 34.6) | 59 | N/A | N/A |
| Poor feeding | 21.7 (16.0 - 27.5) | 192 | 30.4 (18.7 - 42.1) | 59 | N/A | N/A |
| Apnoea/bradycardia | 37.1 (30.5 - 43.8) | 197 | 22.0 (11.4 - 32.6) | 59 | N/A | N/A |

## Discussion

This is the first multinational neonatal stroke active surveillance study using a new purpose-built data platform and standard BPSU approach with either neuroimages or neuroimaging reports reviewed by the research team. The study investigated the incidence, clinical presentation and management of neonatal stroke in the United Kingdom and the Republic of Ireland. The study findings were then compared with previously published active surveillance studies. Unlike our study, previous neonatal stroke active surveillance studies did not collect neuroimages due to the burden of data collection on reporting clinicians, nor did the previous studies report the clinical management and short-term outcomes.

### Incidence

#### Neonatal stroke

The incidence of neonatal stroke was 9·0 cases per 100,000 live births (95% CI 6·9 to 11·6 per 100,000 live births). By definition, as fewer than 1 in 2000 people are affected, neonatal stroke is a rare disease^24^. Less than five cases were included after extending the age range in the case definition to 90 days, rather than 28 days, to capture infants who presented late despite investigations suggesting neonatal stroke. This has not altered the reported incidence rate, with an overall incidence of 8·2 cases per 100,000 live births (95% CI of 6·2 to 10·7) after excluding cases presenting after 28 days of age and preterm infants, in line with previous studies^1^, especially active surveillance studies (**Figure 3**). Our reported neonatal stroke incidence is lower than the incidence of 11 to 15 per 100,000 live births found in a UK-based hospital database study^25^ of neonatal stroke cases presented to neonatal units in England only between 2012 to 2015.

#### Neonatal stroke subtypes

Similar to previous studies^1^, three-quarters of the neonatal stroke cases seen were arterial ischaemic strokes and unilateral, followed by CVST, seen in one in six of the cases. A previous meta-analysis^4^ of 11 published studies and five hospital database records found the incidence of neonatal acute ischaemic stroke in the first 28 days to be 24·6 per 100,000 live births (95% CI 26·7 to 30·4). However, there was a huge variability of the reported neonatal stroke incidence, ranging from 6·1 to 47·3 per 100,000 live births (I^2^=98%). The pooled incidence determined from the five hospital database studies was also significantly higher than that of the 11 literature records (29·3 vs 10·6 per 100,000 live births). When compared with previous surveillance studies (**Figure 3**), the reported incidences of the neonatal stroke subtypes were similar to those found in our study, except for haemorrhagic stroke, where our study reported a lower incidence. This may be due to under-reporting of cases in our study, especially due to the non-specific presentation at a later age.

#### COVID-19 pandemic

The incidence before and after the COVID-19 pandemic appeared similar across the active surveillance studies (**Figure 3**). Data were not collected on maternal COVID-19 infection or vaccination, so further analysis was not possible. However, it was a national recommendation^26^ to offer COVID-19 vaccinations to pregnant women during the current study period. Hence, this does support the absence of adverse neurological findings from the recent Norway/Sweden registry following maternal COVID-19 vaccination^27^ and our previous BPSU survey of neonatal COVID-19 infection^28^.

In summary, our reported neonatal stroke incidence was consistent with the previously published surveillance studies but lower than that reported in hospital database studies. This may partly be explained by the potential error in entering neonatal stroke diagnosis in hospital records, often by non-clinical staff or the most junior member of the clinical team, due to the challenges in diagnosing neonatal stroke. Alternatively, there may be under-reporting of cases in the surveillance study.

### Clinical presentation

The clinical presentation of neonatal stroke is consistent with previously published studies^1,19^. The majority of the neonatal stroke cases were seen in term male White infants presenting in the first two days of age. There is a difference in the clinical presentation of the different neonatal stroke subtypes. The most common presentation for arterial ischaemic and CVST stroke was seizures, while haemorrhagic stroke commonly presented with an encephalopathy picture of altered consciousness and poor feeding, perhaps reflecting the later age at presentation.

Neonatal stroke cases were associated with evidence of fetal distress or hypoxia *in utero*^1^ (**Table 4, Table S2**), with 61% of cases having a cord pH below 7·25, 28% requiring significant resuscitation at birth, and 21% reported as having meconium-stained liquor. 40% of infants with arterial ischaemic stroke were born via emergency Caesarean section. This was nearly twice the emergency Caesarean section rate nationally at that time^29^. This finding was in line with previous studies^1,21^ where infants with arterial ischaemic stroke were commonly delivered by emergency Caesarean section, perhaps raising an interesting question as to whether the fetal distress seen predisposes infants to Caesarean section and higher risk of neonatal stroke, or whether the neonatal stroke led to the fetal distress. On the other hand, most infants with CVST and haemorrhagic stroke were born vaginally. Nearly one-fifth of infants with neonatal stroke have associated congenital cardiac defects, with less than five infants undergoing extracorporeal cardiac membrane oxygenation for transposition of great arteries.

### Clinical management

In accordance with the presenting symptoms, most infants with arterial ischaemic and CVST stroke had an electrocardiogram and cerebral function monitoring with antiseizure treatment. Only one in six infants with neonatal stroke received aspirin or anticoagulant, with over a third of infants with CVST stroke receiving anticoagulant. The majority of infants were discharged home at a median of 12 days old with antiseizure medications alongside paediatric/neonatal and physiotherapy follow-up. Only three-fifths of infants had paediatric neurology follow-up.

The wide variation seen in the investigation, treatment and follow-up provided may be a reflection of the lack of understanding and clinical studies in neonatal stroke. Neonatal stroke guidelines were only available in a quarter of the hospitals which reported neonatal stroke cases. Although a national childhood stroke guideline is available in the UK^30^, there is no national neonatal stroke guideline, despite the higher incidence of stroke in the neonatal period. Furthermore, there is a disparity in the amount of guidance and evidence in a recent scientific statement from the American Heart Association/American Stroke Association^7^ between neonatal and childhood stroke, with more than ten times the amount of references quoted for childhood stroke when compared to neonatal stroke (444 versus 43 references).

### Limitations

There may be under-reporting of neonatal stroke cases, as the study was the first study launched on the dedicated data platform, with a 68% response rate to the BPSU reporting card. However, the standard BPSU active surveillance approach was adopted with the publicity of the study via national neonatal and paediatric neurological associations. Major paediatric centres across the United Kingdom which did not report a neonatal stroke case were contacted to confirm that there were no cases seen during the surveillance period. Although 21% of reported potential cases were unable to be followed up, the actual loss to follow-up cases was likely to be 16% as four cases were likely to be duplicate reports notified by the same centre in the same reporting month, and two cases were likely to be reported outside the surveillance period. The lost to follow-up rate found in our study is consistent with other surveillance studies using similar methodology^9^.

For 20% of reported cases, the case was included in the study based on the clinical information entered by the reporting clinicians, as no neuroimages or reports were provided. However, a consensus meeting with the research group was held to discuss unclear cases. The incidence of neonatal stroke would remain similar at 9·3 cases per 100,000 live births (95% CI 7·1 to 11·9) if the two excluded unclear cases were included in the case definition (**Table S3**).

## Conclusion

This study reported the incidence of neonatal stroke, including its subtypes, in the UK. To the best of our knowledge, this was the first active surveillance study to report the clinical presentation and management of neonatal stroke in the UK and ROI. The study highlighted the differences in the clinical presentation of the different neonatal stroke subtypes and the variation in the management of neonatal stroke. The meta-analysis of the active surveillance studies will improve our understanding of the burden and presentation of neonatal stroke. A standardised approach is needed in investigating and managing infants with neonatal stroke to ensure they receive the best care regardless of where they were born. Further prospective studies, preferably multinational, are needed to fully explore the associated conditions found in this study and elucidate the aetiology of neonatal stroke, including the genomic analyses, which is now a priority in the UK^24^.

## Data Availability

Access to the de-identified data could be requested from the corresponding author after relevant regulatory approvals. The full study protocol, including the study questionnaires, could be found online.

https://www-degruyterbrill-com.nottingham.idm.oclc.org/document/doi/10.1515/med-2022-0554/html

## Acknowledgments

We would like to thank all parents and infants with neonatal stroke participating in the study, as well as clinicians within the UK/Irish Neonatal Stroke Study Collaborative. Please see **Table S4** for the full affiliations of the authors in the collaborative.

## Data Sharing

Access to the de-identified data could be requested from the corresponding author after relevant regulatory approvals. The full study protocol, including the study questionnaires, could be found online.^8^

## References

1. Dunbar M, Kirton A. Perinatal stroke: mechanisms, management, and outcomes of early cerebrovascular brain injury. Lancet Child & Adolescent Health 2018; 2(9): 666–76.

2. Gardner MA, Hills NK, Sidney S, Johnston SC, Fullerton HJ. The 5-year direct medical cost of neonatal and childhood stroke in a population-based cohort. Neurology 2010; 74(5): 372–8.

3. Bemister TB, Brooks BL, Dyck RH, Kirton A. Parent and family impact of raising a child with perinatal stroke. BMC Pediatr 2014; 14: 182.

4. Oleske DM, Cheng X, Jeong A, Arndt TJ. Pediatric Acute Ischemic Stroke by Age-Group: A Systematic Review and Meta-Analysis of Published Studies and Hospitalization Records. Neuroepidemiology. Switzerland: The Author(s). Published by S. Karger AG, Basel.; 2021: 331–41.

5. Hart AR, Connolly DJA, Singh R. Perinatal arterial ischaemic stroke in term babies. Paediatrics and Child Health 2018; 28(9): 417–23.

6. Walther M, Juenger H, Kuhnke N, et al. Motor cortex plasticity in ischemic perinatal stroke: a transcranial magnetic stimulation and functional MRI study. Pediatr Neurol 2009; 41(3): 171–8.

7. Ferriero DM, Fullerton HJ, Bernard TJ, et al. Management of Stroke in Neonates and Children: A Scientific Statement From the American Heart Association/American Stroke Association. Stroke 2019; 50(3): E51–E96.

8. Kwok TC, Dineen RA, Whitehouse W, Lynn RM, McSweeney N, Sharkey D. Neonatal stroke surveillance study protocol in the United Kingdom and Republic of Ireland. Open Med (Wars) 2022; 17(1): 1417–24.

9. Knowles RL, Friend H, Lynn R, et al. Surveillance of rare diseases: a public health evaluation of the British Paediatric Surveillance Unit. Journal of Public Health 2012; 34(2): 279–86.

10. von Elm E, Altman DG, Egger M, et al. The Strengthening the Reporting of Observational Studies in Epidemiology (STROBE) Statement: guidelines for reporting observational studies. Int J Surg 2014; 12(12): 1495–9.

11. Raju TN, Nelson KB, Ferriero D, Lynch JK, Participants N-NPSW. Ischemic perinatal stroke: summary of a workshop sponsored by the National Institute of Child Health and Human Development and the National Institute of Neurological Disorders and Stroke. Pediatrics 2007; 120(3): 609–16.

12. Sébire G, Fullerton H, Riou E, deVeber G. Toward the definition of cerebral arteriopathies of childhood. Curr Opin Pediatr 2004; 16(6): 617–22.

13. Office for National Statistics. Live births by month of occurrence, sex and area of usual residence of mother, England and Wales, September 2021 to August 2023. April 19, 2024. https://www.ons.gov.uk/peoplepopulationandcommunity/birthsdeathsandmarriages/livebirths/adhocs/1973livebirthsbymonthofoccurrencesexandareaofusualresidenceofmotherenglandandwalesseptember2021toaugust2023 (accessed 09/10/2024).

14. National Records of Scotland. Birth Time Series Data. July 30, 2024. https://www.nrscotland.gov.uk/statistics-and-data/statistics/statistics-by-theme/vital-events/births/births-time-series-data (accessed 09/10/2024).

15. Northern Ireland Statistics and Research Agency. Monthly Births. June 11, 2024. https://www.nisra.gov.uk/publications/monthly-births (accessed 09/10/2024).

16. Office for National Statistics. Ethnicity and National Identity in England and Wales: 2011. December 11, 2012. https://www.ons.gov.uk/peoplepopulationandcommunity/culturalidentity/ethnicity/articles/ethnicityandnationalidentityinenglandandwales/2012-12-11 (accessed 24/07/2025).

17. Nyaga VN, Arbyn M, Aerts M. Metaprop: a Stata command to perform meta-analysis of binomial data. Archives of Public Health 2014; 72(1): 39.

18. Sorg A-L, von Kries R, Klemme M, et al. Risk factors for perinatal arterial ischaemic stroke: a large case–control study. Developmental Medicine & Child Neurology 2020; 62(4): 513–20.

19. Sorg A-L, Klemme M, von Kries R, et al. Clinical Diversity of Cerebral Sinovenous Thrombosis and Arterial Ischaemic Stroke in the Neonate: A Surveillance Study. Neonatology 2021; 118(5): 530–6.

20. Klemme M, Gerstl L, Weinberger R, et al. Neonatal Arterial Ischemic Stroke - A Hospital Based Active Surveillance Study in Germany. Klin Padiatr 2017; 229(3): 142–6.

21. Roy B, Webb A, Walker K, et al. Prevalence & Risk Factors for Perinatal Stroke: A Population-Based Study. Child Neurol Open 2023; 10: 2329048x231217691.

22. Roy B, Webb A, Walker K, Morgan C, Badawi N, Novak I. Risk factors for perinatal stroke in term infants: A case–control study in Australia. Journal of Paediatrics and Child Health 2023; 59(4): 673–9.

23. Statistisches Bundesamt Deutschland. Live births and difference to the previous year. July 1, 2025. https://www.destatis.de/EN/Themes/Society-Environment/Population/Births/Tables/live-birth-difference.html (accessed 01/08/2025).

24. The UK Rare Diseases Framework. January 9, 2021. https://www.gov.uk/government/publications/uk-rare-diseases-framework/the-uk-rare-diseases-framework#priorities (accessed 15/07/2025).

25. Gale C, Jeyakumaran D, Ougham K, Jawad S, Uthaya S, Modi N. Brain injury occurring during or soon after birth: annual incidence and rates of brain injuries to monitor progress against the national maternity ambition 2016 and 2017 data. London: National Data Analysis Unit, Imperial College London, 2019.

26. Royal College of Obstetricians and Gynaecologists (RCOG). Coronavirus (COVID-19) Infection in Pregnancy. December, 2022. https://www.rcog.org.uk/guidance/coronavirus-covid-19-pregnancy-and-women-s-health/coronavirus-covid-19-infection-in-pregnancy (accessed 19/08/2025).

27. Norman M, Magnus MC, Söderling J, et al. Neonatal Outcomes After COVID-19 Vaccination in Pregnancy. JAMA 2024; 331(5): 396–407.

28. Gale C, Quigley MA, Placzek A, et al. Characteristics and outcomes of neonatal SARS-CoV-2 infection in the UK: a prospective national cohort study using active surveillance. Lancet Child Adolesc Health 2020.

29. NHS Digital. NHS Maternity Statistics, 2022-23 December 7, 2023. https://digital.nhs.uk/data-and-information/publications/statistical/nhs-maternity-statistics/2022-23 (accessed 15/07/2025).

30. Royal College of Paediatrics and Child Health. Stroke in childhood - Clinical guideline for diagnosis, management and rehabilitation. May 2017. https://www.rcpch.ac.uk/sites/default/files/2024-12/2017_Stroke_in_childhood_guideline_final.pdf.pdf (accessed 01/08/2025).

